# Management Of Rural Acute Coronary Syndromes (MORACS) – A randomized controlled trial of diagnostic support for patients in rural hospitals with acute coronary syndromes

**DOI:** 10.1101/2025.11.26.25341123

**Authors:** Matthew French, Georgia Meagher, Christopher Ray Chen, Fiona Dee, Lindsay Savage, James W. Leitch, Conrad Loten, Olivia Watson, Kerry J. Inder, Dawn McIvor, Trent Williams, Peter Fletcher, John French, Ammar Albayati, Shanathan Sritharan, Daniel Barker, Ashleigh Stuart, Mohammed S Al-Omary, Allan J. Davies, John Attia, John Wiggers, Aaron L. Sverdlov, Nicholas Collins, Andrew J. Boyle

## Abstract

**Background:** Rural and remote patients presenting with acute coronary syndrome (ACS) experience delays in diagnosis and reperfusion, contributing to higher mortality than those presenting to tertiary centres. Evidence supporting system-triggered diagnostic interventions across the full spectrum of ACS remains limited.

**Methods:** In a multicentre, cluster-randomized controlled trial, 29 rural emergency departments in New South Wales, Australia, were assigned to either a system-triggered diagnostic telemedical support from a tertiary centre (MORACS intervention) or usual care. Adult patients presenting with symptoms consistent with ACS were included in the study. The primary outcome was all-cause death. Secondary outcomes included 30-day and 1-year death, length of hospital stay and 30-day readmission rate.

**Results:** Between December 2018 and April 2020, ACS was confirmed in 587 of 7474 emergency department presentations consistent with possible ACS. Of these, 274 (47%) presented to MORACS intervention hospitals and 313 (53%) presented to control hospitals. Over a median follow-up of 62 months, all-cause death occurred in 54 patients (20%) in the intervention group and 85 patients (27%) in the control group, representing a 30% lower risk of death with the MORACS intervention (HR 0.70; 95% CI, 0.49–0.98; P = 0.036). The mortality benefit was consistent across both the ST-elevation ACS (STEACS) and non-ST elevation ACS (NSTEACS) subgroups. No significant differences were observed in length of stay or 30-day readmissions.

**Conclusions:** The MORACS intervention significantly reduced all-cause mortality among rural patients with ACS. These findings support the integration of system-triggered diagnostic telehealth support into rural emergency department settings to improve outcomes in ACS management.

**Registration:** ACTRN12619000533190 (anzctr.org.au)

**Clinical Perspective:** *What is New?:*

- In this multicentre cluster-randomized controlled trial, system-triggered diagnostic telemedical support in rural emergency departments significantly reduced all-cause mortality in patients presenting with ACS.
- This larger cohort extends the findings of the initial MORACs study, demonstrating a mortality benefit of diagnostic telemedical support across the full spectrum of ACS presentations, including both STEACS and NSTEACS.

*What Are the Clinical Implications?:*

- Implementing system-triggered diagnostic telemedical support can help overcome barriers to timely diagnosis and reperfusion in geographically isolated hospitals.
- Broader implementation of such interventions may enhance equity in cardiac care delivery and reduce rural-urban disparities in ACS outcomes.

## Introduction

Acute coronary syndrome (ACS) remains a leading cause of morbidity and mortality worldwide, responsible for more than 15 million hospitalisations and 9 million deaths (approximately 16% of all global deaths) annually^1^. Outcomes are dependent on rapid and accurate diagnosis and initiation of reperfusion or revascularisation therapy. For ST-elevation ACS (STEACS), international guidelines recommend primary percutaneous intervention (PCI) within 90 minutes of first medical contact or fibrinolysis followed by urgent transfer if PCI delay is expected^2^. For non-ST elevation ACS (NSTEACs), early risk stratification and angiography within 2-72 hours is also guideline directed standard of care^2^.

Rural and remote patients experience poorer outcomes due to delays in diagnosis and reperfusion (geographic distance to PCI-capable centres, limited access to medical specialists), and higher cardiovascular risk burden^3,4^. In Australia, approximately 30% of the population live in rural and remote areas. New South Wales, Australia’s most populated state, spans over 800 000 km ² (approximately 309,000 square miles - 18% larger than Texas) and public health care is organized into geographically defined Local Health Districts (LHD). Many small hospitals are staffed by general practitioners or career medical officers without emergency medicine or cardiology expertise. Within the Hunter New England Local Health District (HNELHD) where this study is conducted, one tertiary hospital provides 24-hour PCI coverage across >130,000 km² (>50,000square miles), serving >1 million people, including a higher than average Indigenous population (5.2-15% versus national 3.2%)^5^.

To improve access to reperfusion, NSW established a statewide Cardiac Reperfusion Strategy (SCRS) enabling prehospital electrocardiograph transmission and remote specialist interpretation (by emergency medicine or cardiology physicians) for ambulance-identified STEACS^6^. However, this pathway does not include the 30% of STEACS who self-present to rural emergency departments or all patients with NSTEACS who remain at high risk of diagnostic delay and adverse outcomes^7, 8^.

The Management of Rural Acute Coronary Syndromes (MORACS) randomized control trial was designed to evaluate whether a system-triggered real-time diagnostic telemedical support service could improve outcomes for patients presenting with ACS to rural hospitals. Earlier evaluation of the MORACS intervention in the STEACS subgroup demonstrated reduced time to fibrinolysis and improved diagnostic accuracy for STEACS diagnosis^9^. The present study extends these findings to the entire ACS spectrum and evaluates mortality outcomes.

## Methods

### Study Design and Setting

The MORACS trial was a prospective, multicentre, cluster-randomized controlled trial evaluating the effectiveness of a system-triggered, real-time diagnostic and treatment telemedical support for Acute Coronary Syndrome (ACS).

The trial was conducted across 29 rural hospitals within the HNELHD in NSW, Australia. Hospitals were eligible if their emergency departments were staffed exclusively by general practitioners/primary care practitioners or career medical officers/hospitalists. Specifically, there were no emergency-medicine specialists (defined as Fellows of the Australasian College for Emergency Medicine [FACEM] or international equivalent) in these hospitals. Two PCI-capable centres served this region: John Hunter Hospital (Newcastle; 24 hour PCI service) and Tamworth Regional Referral Hospital (Tamworth; limited PCI availability during business hours). Eligible hospitals ranged from first-aid posts without inpatient capacity to hospitals with 100 inpatient beds and staffing consistent of both general practitioners and career medical offers and some hospitals were >650 km from a PCI centre.

The trial adhered to the Declaration of Helsinki and the CONSORT 2025 reporting guidelines^10^. Ethics approval was obtained from the Hunter New England Health Service Human Research Ethics Committee. Cluster-level consent was obtained from hospital executives.

### Randomisation and Masking

Randomisation occurred at the hospital level, stratified by hospital size and staffing model (general practitioners vs career medical officers). Fifteen hospitals were assigned to the MORACS intervention and fourteen to usual care. Outcomes assessment was blinded to group allocation.

### Participants

Eligible participants were adults presenting to participating hospitals between December 2018 and April 2020 with symptoms suggestive of ACS (defined by ICD-10 codes; Supplement Table 1). Exclusion criteria included age < 30 years (< 20 years for Indigenous patients), prehospital STEACS diagnosis with fibrinolysis via ambulance pathway, traumatic cardiac arrest without return of spontaneous circulation, non-cardiac trauma, or palliative care management. This analysis extends the original MORACS STEACS trial (ACTRN12619000533190) to include all ACS subtypes.

### Study Procedures

At both intervention and control hospitals, a patient triaged with an ICD-10 code compatible with ACS initiated an automatic text message notification to one of three MORACS specialist cardiac nurses at the tertiary referral centre. The MORACS nurse had real-time access to each patients ECG and point of care troponin via the electronic medical record system and adjudicated all presentations as either STEACS, NSTEACS or non-ACS. STEACS was defined using ECG and clinical presentation criteria from the fourth universal definition of myocardial infarction^11^. NSTEACS was defined using ECG and clinical presentation criteria and an elevated serum Troponin-I assay (Abbott Architect STAT i2000SR hs-cTnI; ≥16 ng/L for females, ≥26 ng/L for males) ^12^. Many of the rural centres only had access to point of care troponin testing (Abbott i-STAT cTnI); in those cases, their first formal Troponin-I was taken after transfer to a centre where the assay was available.

At intervention hospitals only, MORACS nurses initiated contact with local treating clinicians to provide diagnostic support and treatment recommendations, including transfer to a PCI-capable facility in accordance with the New South Wales Cardiac Reperfusion Strategy (SCRS) and the associated Pathway for Acute Coronary Syndromes in Adults (PACSA) guideline^16^. Local clinicians could also initiate contact with the MORACS team, though activation of the intervention did not require an “opt-in” step because contact was system-triggered. The intervention operated between 9:00 AM and 7:30 PM seven days per week with one nurse rostered per day. When patients presented outside of MORACS hours, they were treated by local clinicians as per the established pathway and reviewed by the MORACS nurse at the commencement of the following shift (9:00 am). The relevant intervention hospital was contacted if the patient remained in the hospital or in the event a patient had been or was being discharged and required further assessment.

Control hospitals followed existing local protocols for the management of suspected ACS and had no automated telemedical alerts or proactive tertiary-centre engagement, although clinicians could seek cardiology advice through standard referral channels.

### Outcomes

The primary outcome was all-cause mortality, determined from the NSW death registry data. Survival time was measured from index presentation to date of death and patients without a recorded death by February 2, 2024 were censored. Secondary outcomes included 30-day and 1-year mortality, index hospital length of stay, and 30-day readmission (representation requiring ≥ 1 overnight stay).

### Statistical Analysis

Statistical analyses were performed using SAS version 9.4 (SAS Institute, Cary, NC). Categorical variables were summarized as number (percentage), and continuous variables as mean ± SD or median (interquartile range), as appropriate. Between-group comparisons used χ² or Fisher exact tests for categorical variables and Wilcoxon or Kruskal–Wallis tests for continuous variables.

All-cause mortality was analyzed using Cox proportional hazards regression to estimate hazard ratios (HRs) and 95% confidence intervals (CIs). The proportional hazards assumption was verified both graphically and using Schoenfeld residuals. Kaplan–Meier survival curves and log-rank tests were used to compare survival distributions. Prespecified subgroup analyses were conducted for ST-elevation ACS (STEACS) and non–ST-elevation ACS (NSTEACS). A two-sided P value < 0.05 was considered statistically significant.

## Results

From December 2018 through to April 2020, there were 7,474 rural hospital emergency department presentations (in 6,249 patients) that had ICD-10 triage codes compatible with ACS. Of these, 587 patients had a final diagnosis consistent with ACS (21% [n=123] STEACS, 79% [n=464] NSTEACS) and met inclusion criteria for the study. 47% of patients (46 STEACS, 228 NSTEACS) presented to hospitals assigned to the MORACS intervention and 53% (77 STEACS, 236 NSTEACS) presented to hospitals assigned to usual care (Figure 1).

**Figure 1.**
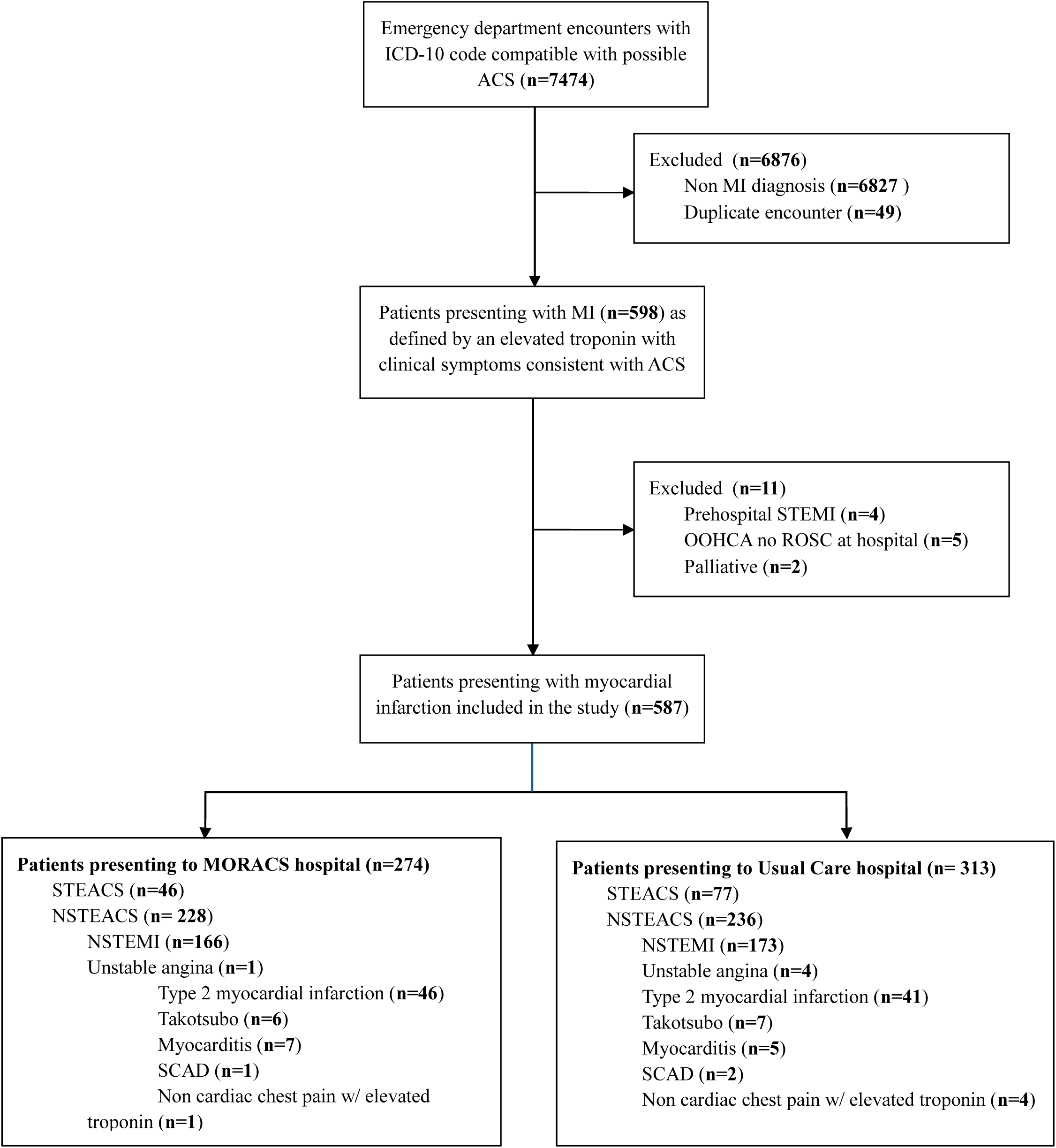
Consort Diagram.

Baseline characteristics of patients are included in Table 1. The mean age of participants was 68 ± 14 years, 67% (n=393) were male and 12% (n=73) identified as Indigenous (Aboriginal or Torres Strait Islander). 36% (n=210) participants had a known history of ischemic heart disease prior to their presentation. Of all comorbidities, hypertension was the most common, present in 64% (n=358) of participants, followed by dyslipidaemia in 41% (n=231) and diabetes in 30% (n=173). Within the NSTEACS cohort, 73% (n=339) of patients had a diagnosis of type 1 myocardial infarction (T1MI) and 19% (n=87) had a type 2 myocardial infarction (T2MI). The diagnosis of the remaining 38 patients is included in Figure 1.

**Table 1.**
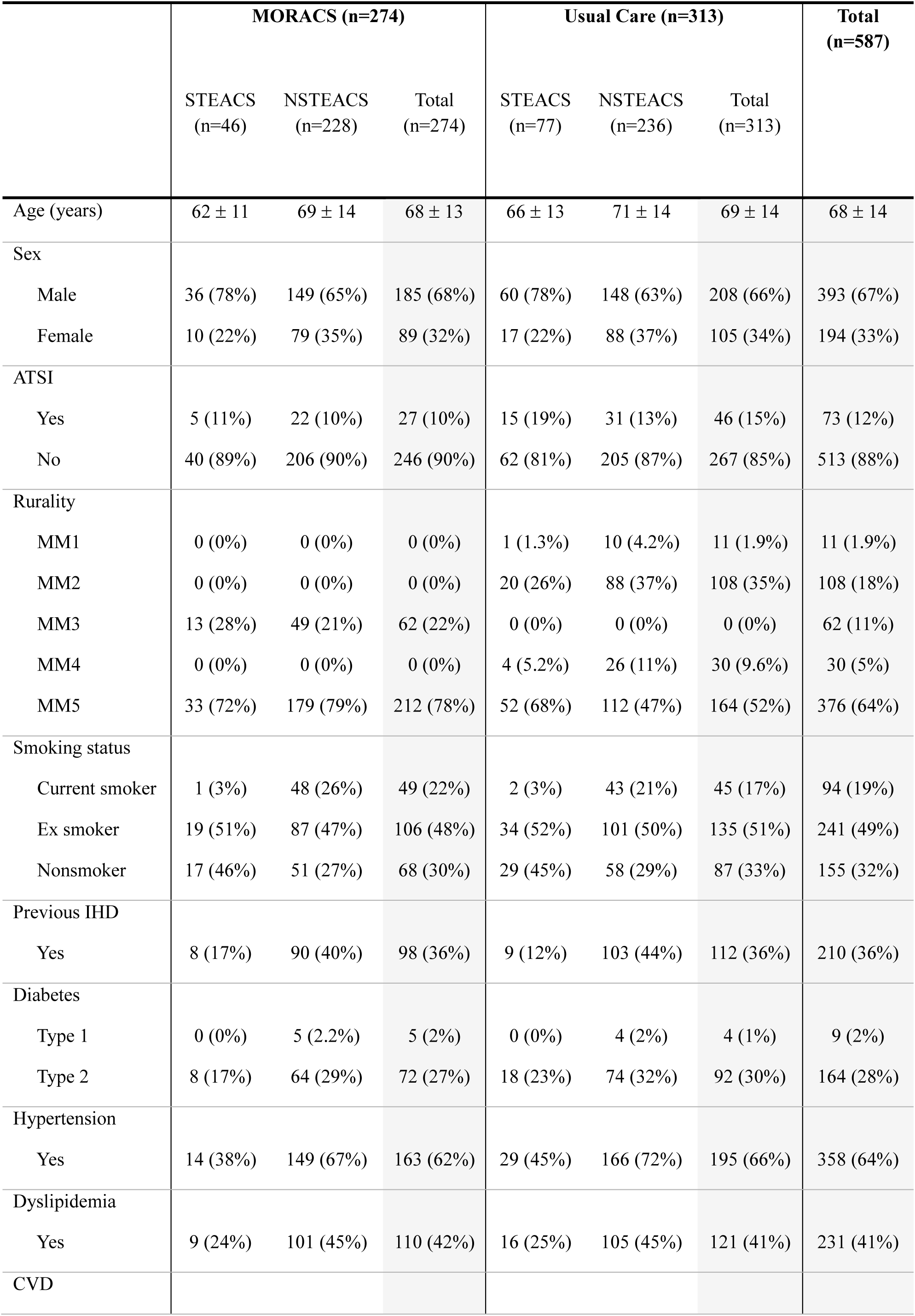
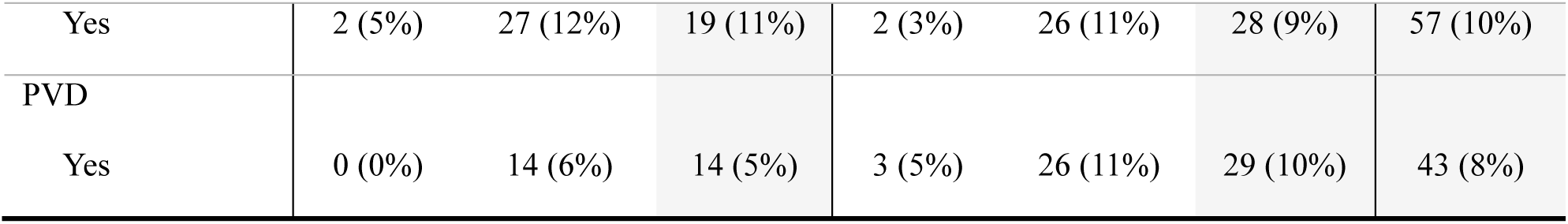
Patient characteristics.

### Primary Outcome

Over a median follow-up of 62 months, there were 54 deaths (19.7%) in the MORACS group and 85 (27.2%) deaths in the usual care group (HR 0.70 [95% CI 0.49 – 0.98], p=0.036) (Figure 2). The Kaplan-Meier survival curves continue to separate over time, suggesting the MORACs intervention resulted in sustained and continued mortality benefits. This trend was observed in both the STEACS and NSTEACS populations (supplemental Figures 2 and 3) and in all clinical subgroups (Figure 3).

**Figure 2.**
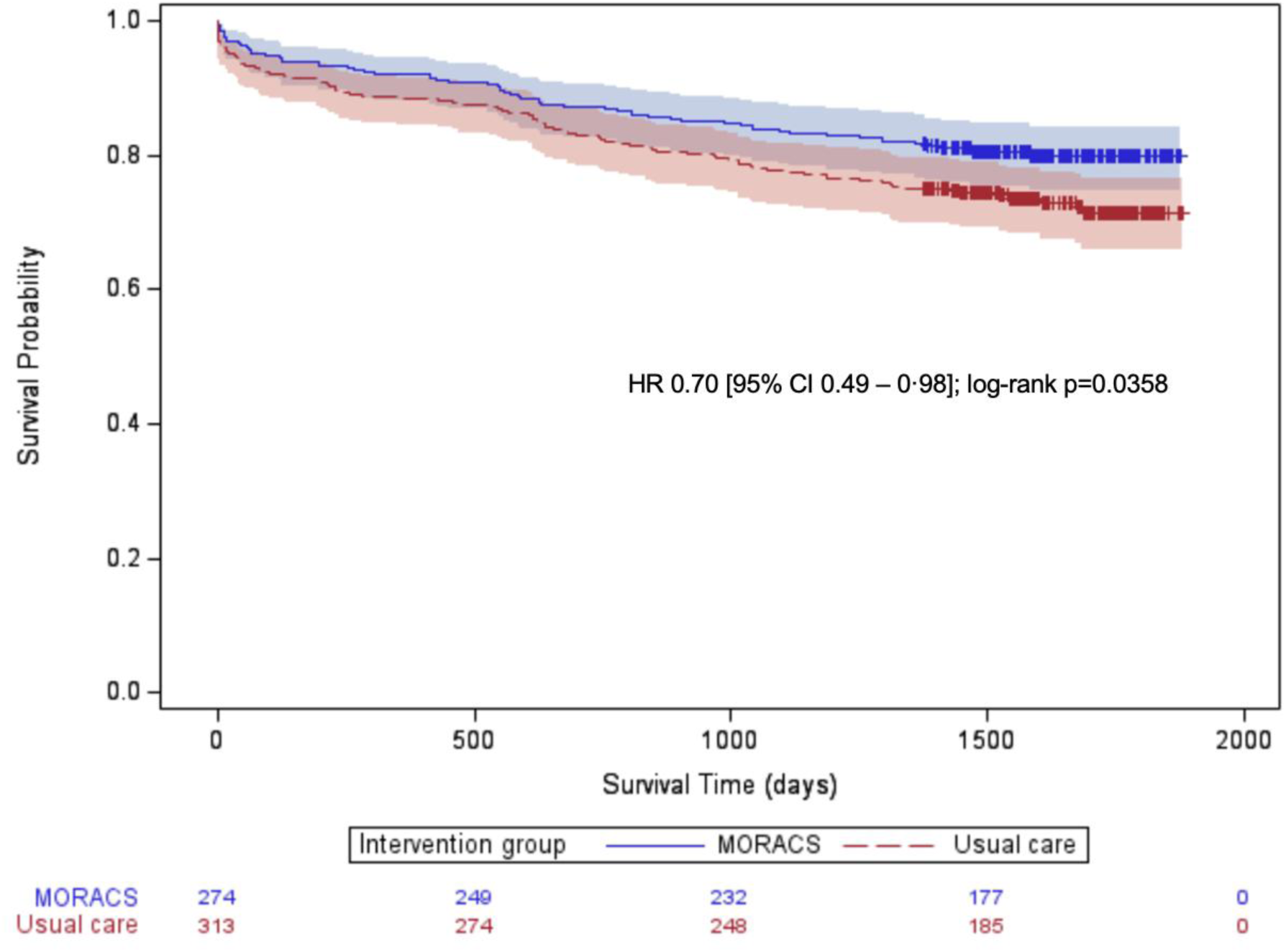
Kaplan-Meier plot for entire cohort with number of subjects at risk and 95% confidence limits.

**Figure 3.**
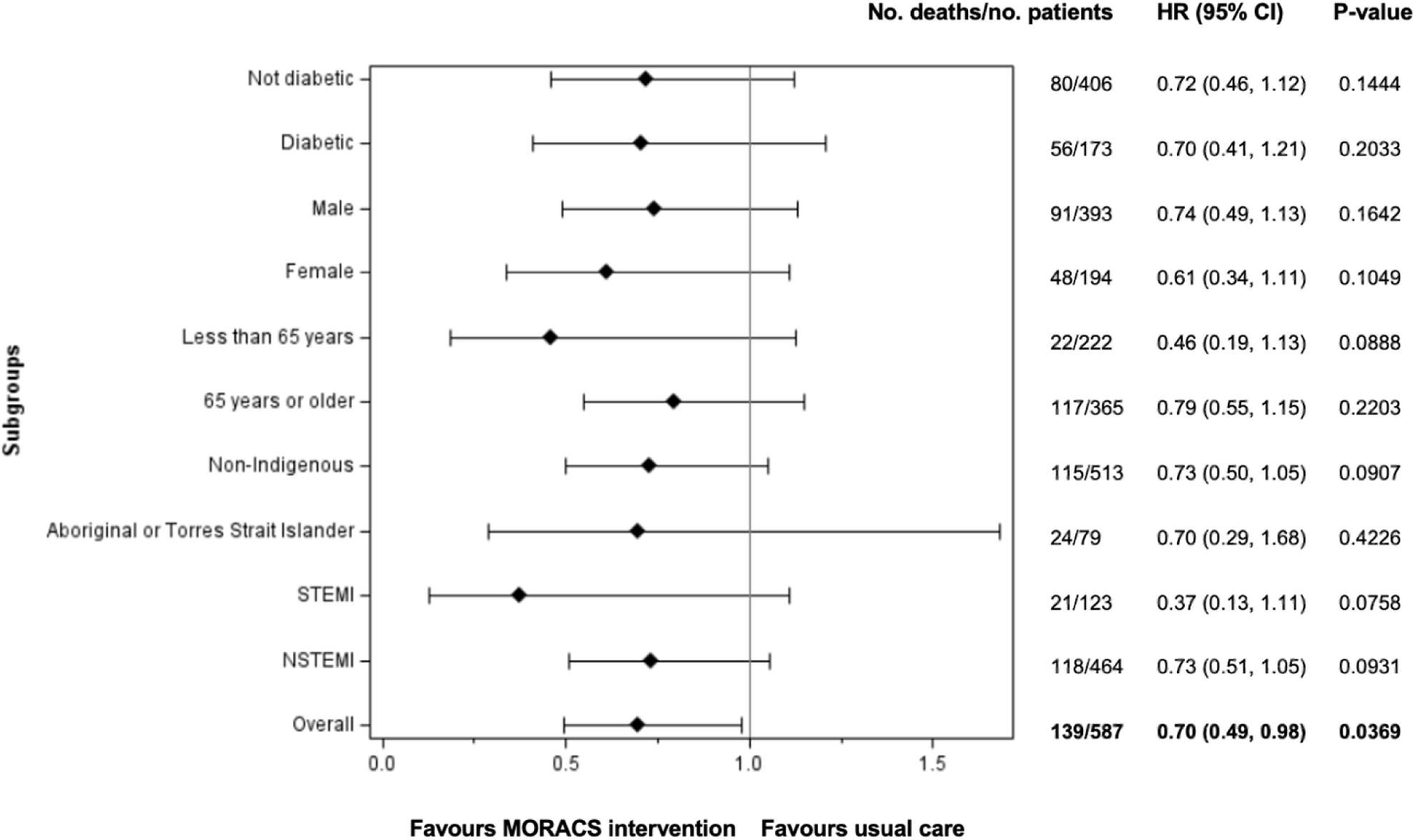
Forest Plot of Subgroups.

### Secondary Outcomes

There was no statistically significant difference in the number of all-cause deaths at 30 days (8 vs. 15 for MORACS vs. usual care; p=0.29) or 1 year (22 vs. 36; p=0.17). There was no difference in overall length of stay during the index admission, which incorporated both the stay at the rural hospital and at the tertiary referral hospital. 30 day readmission was not significantly different between the intervention and control group. (Table 2)

**Table 2.**
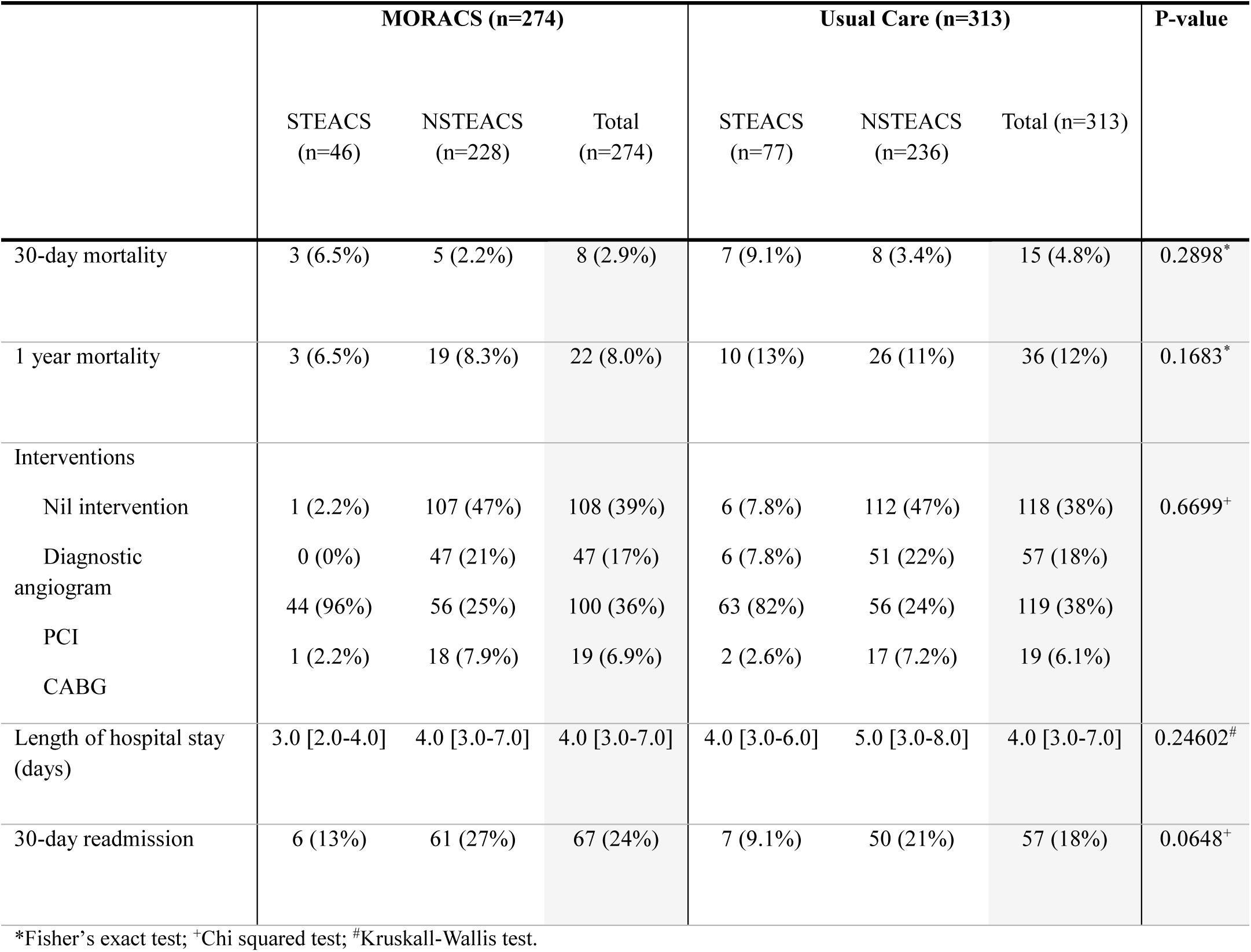
Secondary Outcomes.

## Discussion

Prompt diagnosis is essential in the management of ACS. International guidelines recommend that a 12-lead ECG be performed within ten minutes of first medical contact.^6^ Rural physicians, however, infrequently encounter STEACS and may lack the diagnostic experience of clinicians in high-volume tertiary cardiac centers. A recognized shortfall in healthcare staffing outside major metropolitan areas further compounds the scarcity of acute cardiac expertise.^15^ This study shows that a diagnostic support service—staffed by nurses with advanced ECG interpretation skills and integrated with point-of-care troponin testing—can bridge this expertise gap and significantly reduce mortality. These findings underscore the value of a targeted, low-cost intervention in addressing inequities in access to high-quality acute cardiac care.

A key strength of this trial is that it focused solely on diagnostic support. Once ACS was identified, patients entered existing treatment pathways, which differ for STEACS and NSTEACS. As such, the intervention did not increase clinician workload, alter inter-hospital transfer protocols, or extend hospital length of stay. Notably, almost all transfers made under this protocol were clinically appropriate (Supplementary Data). This aligns with previous findings^8^ that delayed or incorrect diagnosis is a major contributor to morbidity and mortality in rural hospitals. The present study demonstrates that improving initial diagnosis through the simple MORACS intervention translates directly into lower mortality.

Length of stay was significantly reduced by the MORACS intervention in the STEACS subgroup (by approximately one day) but not in the overall cohort. The majority of participants presented with NSTEACS rather than STEACS. This older, multimorbid NSTEACS population typically requires longer hospitalizations, which likely diluted the effect seen in the STEACS subgroup.

Current models of care for regional hospitals in Australia generally rely on an “outside-in” escalation of care: regional centers follow guideline-based referral pathways for ACS^16^ and depend on early recognition by peripheral healthcare providers. The MORACS intervention represents an “inside-out” model, allowing experienced tertiary-center clinicians to extend diagnostic oversight to resource-and experience-limited peripheral hospitals. In this model, a simple text message alert can initiate expert review and potentially save lives.^17^

This diagnostic support framework is scalable and adaptable across diverse healthcare environments, particularly those facing similar workforce shortages and geographic or resource constraints. Future work should include formal economic evaluation to determine cost-effectiveness, given the promising clinical outcomes observed. Additionally, automating ECG machine integration with electronic medical records could further enhance system responsiveness and accessibility.

Another strength of this study is its robust design—a large, randomized trial clustered by hospital size and conducted across a vast and demographically diverse geographic region, improving generalizability to many rural contexts. However, results may not be directly transferable to all healthcare systems, funding structures, or cultural settings. Due to cost limitations, the intervention did not provide full 24-hour coverage. Future implementation should explore strategies for continuous, after-hours operation to maximize benefit.

## Conclusion

The MORACS trial showed that a system-triggered diagnostic support system can reduce mortality in patients presenting with acute coronary syndromes to small rural hospitals.

## Data Availability

De-identified individual participant data (including data dictionaries) that underlie the results reported in this article will be made available to researchers who provide a methodologically sound proposal. Proposals should be directed to the corresponding author; to gain access, data requestors will need to sign a data access agreement. Data will be available beginning 6 months and ending 5 years following article publication.

## Acknowledgments

AB and FD were responsible for conceptualisation of the MORACS intervention. AS and AB were responsible for funding acquisition. FD, AB and MF contributed significantly to the study conception and design. MF, GM, AB and MA analysed and interpreted the data. MF, GM, MA, AB, DB and AS conducted the statistical analysis. MF and GM wrote the manuscript. AB supervised the work of all other authors. All other authors contributed to the implementation and data collection. All authors were involved in the data interpretation and discussion of the results. All authors critically revised the manuscript and approved the final version for submission. All authors had full access to all data in the study and had final responsibility for the decision to submit for publication.

## References

1. Jneid H, Addison D, Bhatt DL, Fonarow GC, Gokak S, Grady KL, et al. 2017 AHA/ACC Clinical Performance and Quality Measures for Adults With ST-Elevation and Non-ST-Elevation Myocardial Infarction: A Report of the American College of Cardiology/American Heart Association Task Force on Performance Measures. Circ Cardiovasc Qual Outcomes. 2017;10(10).

2. Brieger D, Cullen L, Briffa T, Zaman S, Scott I, Papendick C, et al. National Heart Foundation of Australia & Cardiac Society of Australia and New Zealand: Comprehensive Australian Clinical Guideline for Diagnosing and Managing Acute Coronary Syndromes 2025. Heart, Lung and Circulation. 2025;34(4):309–97.

3. Health AIo, Welfare. Rural and remote health. Canberra: AIHW; 2024.

4. Khan AA, Williams T, Savage L, Stewart P, Ashraf A, Davies AJ, et al. Pre-hospital thrombolysis in ST-segment elevation myocardial infarction: a regional Australian experience. Medical Journal of Australia. 2016;205(3):121–5.

5. Health: NMo. Centre for Epidemiology and Evidence. HealthStats NSW. Sydney: NSW Ministry of Health: Australian Bureau of Statistic; 2021 [Available from: https://www.healthstats.nsw.gov.au/r/120637.

6. Innovation AfC. State Cardiac Reperfusion Strategy 2025 [Available from: https://aci.health.nsw.gov.au/networks/cardiac/resources/scrs.

7. Chew DP, French J, Briffa TG, Hammett CJ, Ellis CJ, Ranasinghe I, et al. Acute coronary syndrome care across Australia and New Zealand: the SNAPSHOT ACS study. Medical Journal of Australia. 2013;199(3):185–91.

8. Williams T, Savage L, Whitehead N, Orvad H, Cummins C, Faddy S, et al. Missed Acute Myocardial Infarction (MAMI) in a rural and regional setting. Int J Cardiol Heart Vasc. 2019;22:177–80.

9. Dee F, Savage L, Leitch JW, Collins N, Loten C, Fletcher P, et al. Management of Acute Coronary Syndromes in Patients in Rural Australia: The MORACS Randomized Clinical Trial. JAMA Cardiology. 2022;7(7):690–8.

10. Hopewell S, Chan A-W, Collins GS, Hróbjartsson A, Moher D, Schulz KF, et al. CONSORT 2025 statement: updated guideline for reporting randomized trials. BMJ. 2025;389:e081123.

11. Thygesen K, Alpert JS, Jaffe AS, Chaitman BR, Bax JJ, Morrow DA, et al. Fourth Universal Definition of Myocardial Infarction (2018). Circulation. 2018;138(20):e618–e51.

12. Apple FS, Ler R, Murakami MM. Determination of 19 cardiac troponin I and T assay 99th percentile values from a common presumably healthy population. Clin Chem. 2012;58(11):1574–81.

13. Boeddinghaus J, Nestelberger T, Koechlin L, Wussler D, Lopez-Ayala P, Walter JE, et al. Early Diagnosis of Myocardial Infarction With Point-of-Care High-Sensitivity Cardiac Troponin I. Journal of the American College of Cardiology. 2020;75(10):1111–24.

14. AIHW, R K, C H, J L. Older Australians in hospital. Canberra: AIHW; 2007.

15. Cortie CH, Garne D, Parker-Newlyn L, Ivers RG, Mullan J, Mansfield KJ, et al. The Australian health workforce: Disproportionate shortfalls in small rural towns. Australian Journal of Rural Health. 2024;32(3):538–46.

16. NSW Government, Agency for Clinical Innovation. Pathway for Acute Coronary Syndrome (PACSA) 2025 [Available from: https://aci.health.nsw.gov.au/networks/eci/clinical/tools/cardiology/pathway-for-acute-coronary-syndrome. Last updated October 2025.

17. Ofoma UR, Joynt Maddox KE. Improving the Detection of ST-Segment Elevation Myocardial Infarction in Rural Settings: When Texting Saves Lives. JAMA Cardiol. 2022;7(7):698–9.

